# Assessing Self-reported and Device-Derived Sleep Quality in a Sample of Older Adults with Chronic Musculoskeletal Pain

**DOI:** 10.1101/2025.03.17.25324131

**Authors:** Soamy Montesino-Goicolea, Pedro Antonio Valdes-Hernandez, Olga Nin, Cameron Smith, Eric C. Porges, Yenisel Cruz-Almeida

**Affiliations:** Pain Research & Intervention Center of Excellence, University of Florida, Gainesville, FL, USA; Department of Community Dentistry & Behavioral Science, College of Dentistry, University of Florida, Gainesville, FL, USA; Center for Cognitive Aging & Memory, McKnight Brain Foundation, University of Florida, Gainesville, FL, USA; Department of Anesthesiology, College of Medicine, University of Florida, Gainesville, FL, USA; Department of Clinical and Health Psychology College of Public Health and Health Professions, University of Florida, Gainesville, FL, USA; Institute on Aging, University of Florida, Gainesville, FL, USA

**Author notes:** First and second authors contributed equally to this work. Corresponding author information: Yenisel Cruz-Almeida, MSPH, PhD, PO Box 103628, 1329 SW 16th Street, Ste 5180 (zip 32608), Gainesville, FL 32610, URL: https://price.ctsi.ufl.edu/about-the-center/staff/yenisel-cruz-almeida/.

**Keywords:** Oura Ring, Subjective Sleep, Objective Sleep, Clinical Pain, Experimental Pain

## Abstract

**Objectives:** Our primary aim was to evaluate the agreement between subjective and objective methods of measuring sleep quality in a musculoskeletal pain sample. Secondly, we aimed to explore the relationship between subjective and objective sleep quality—and its impact on function—and clinical and experimental pain.

**Methods:** We assessed subjective sleep using the Pittsburgh Sleep Quality Index (PSQI) and objective sleep using the Oura ring—a wearable characterizing sleep stages. Participants had musculoskeletal pain (intensity>4/10 most days in past 3 months) and poor sleep (PSQI total>5). To enable direct comparisons, via correlations, between subjective and objective sleep (primary aim), we emulated the equivalent of PSQI’s answers and components by averaging the appropriate Oura data over the month covered by the PSQI. We used partial correlations to assess sleep-pain relationships (second aim)—controlling for age and sex.

**Results:** Answers to PSQI questions about total bedtime and sleep duration, and the PSQI duration component, correlated with their Oura equivalents, whereas PSQI failed to capture Oura’s Sleep Latency, Efficiency, and Disturbances. On the other hand, PSQI total score and its sleep latency component correlated with WOMAC-pain score, MPQ scores (total, neuropathic, continuous, and intermittent) and GCPS-pain intensity, while Oura’s Sleep Latency correlated with conditioned pain modulation. No significant association between Oura measures and pain was found.

**Conclusions:** The findings highlight the complementary roles of subjective and objective measures and the need for integrated approaches to refine sleep assessments in musculoskeletal pain. Future studies should investigate the causes of these discrepancies to enhance understanding of sleep-related health outcomes.

## Introduction

Sleep quality and pain are two critical aspects of health that significantly influence the well-being and daily functioning of middle-aged and older adults^19^. Chronic pain, prevalent in aging populations, is closely linked with sleep disturbances in a cycle where each exacerbates the other^37^. Poor sleep heightens pain sensitivity, while chronic pain disrupts sleep architecture, reducing sleep quality and complicating pain management^3,19,22,27^. Consequently, accurately assessing sleep quality in this population is essential for developing interventions to mitigate both pain and sleep disturbances, thereby improving quality of life ^13^.

The Pittsburgh Sleep Quality Index (PSQI)^7^ is a widely recognized and validated gold-standard for assessing subjective self-reported sleep quality, known for its broad applicability across clinical and non-clinical populations and its frequent use as a reference measure in psychometric research^18^. It assesses various aspects of sleep, including duration, latency, disturbances, and overall quality over the past month. While valuable in capturing individuals’ perceptions of their sleep, the PSQI is prone to recall bias and personal perceptions, which may lead to discrepancies between reported and actual sleep experiences.

Wearable technology has made objective measures of sleep quality more accessible. Devices like the Oura ring (OURA©) use infrared sensors and accelerometers to track sleep stages, providing a comprehensive view of sleep patterns^16,32^, measuring parameters like sleep latency, total sleep duration, sleep efficiency, and heart rate variability^2^. Moreover, they are sensible to aspects of sleep often overlooked or inaccurately reported in self-assessments, such as micro-arousals and subtle disruptions^16,32^. The non-intrusive design and ease of use of the Oura ring make it particularly suited for older adults, a population in which chronic pain is more prevalent, as it promotes compliance over extended periods.

Although subjective and objective measures assess the same construct (i.e., sleep quality) discrepancies may arise, particularly in metrics like sleep latency and efficiency, where subjective perceptions may diverge from objective measures (e.g., PQSI versus polysomnography or actigraphy^7^). These discrepancies are especially relevant in chronic pain populations, where subjectivity is influenced by the multidimensional experience of pain within its biopsychosocial framework^9,21,31^. Understanding these divergences not only sheds light on the perceptual biases of chronic pain patients regarding their sleep disturbances but also informs the development of targeted interventions that address sleep quality in individuals with chronic pain.

This study primarily aimed to bridge the gap between subjective and objective sleep assessments in older adults with musculoskeletal pain by comparing PSQI responses and components with their equivalents estimated from Oura ring data, where applicable. We hypothesized that PSQI measures would significantly and positively correlate with their Oura-based equivalents. The expectation of a positive correlation is based on the rationale that, by design, our Oura equivalents are supposed to mirror the PSQI metrics. Only one study has reported significant correlations between PSQI and Oura data^12^—PSQI total score significantly correlated with Oura’s proprietary Sleep Duration and Efficiency—without tailoring the Oura data to align with PSQI metrics.

Additionally, we explored the relationship between sleep and pain in our sample. We hypothesized that subjective sleep-related measures—derived from the PSQI and other questionnaires assessing the impact of sleep quality on function (e.g., daytime sleepiness, daily living activities, and impairment)—would significantly correlate with clinical or experimental pain. Furthermore, we hypothesized that Oura-derived objective sleep quality measures would also significantly correlate with clinical or experimental pain.

By integrating subjective and objective assessments, this study highlights the importance of a multimodal approach to sleep evaluation in clinical practice. Such an approach may offer a more comprehensive understanding of the sleep-pain relationship and supports the development of personalized interventions to enhance sleep quality and health outcomes for individuals with chronic pain.

## Participants and Methods

### Ethics statement

The study was approved by the University of Florida (UF) Institutional Review Boards and carried out in accordance with the Declaration of Helsinki. Participants provided verbal and written informed consent.

### Participants

This is a secondary data analysis of a pilot study double-blinded, placebo-controlled, randomized parallel group clinical trial designed to examine the feasibility of a four-week regimen of oral Aminobutyric Acid or γ-aminobutyric acid (GABA) intake^29^. The total sample consisted of 33 middle-aged individuals recruited between March 2021 and February 2023. Inclusion eligibility of the parent study was determined through telephone screening.

A detailed description of the screening, inclusion, and exclusion criteria of the parent study was reported previously^30^. In brief, older adults 45 years of age and older who had a smartphone, pain of at least moderate intensity (>5/10 rating) on more days than not during the past three months, and poor sleep quality (PSQI Total>5) were considered for participation. Participants were excluded due to inability to consent, pregnancy, cognitive impairment, psychiatric and certain neurological conditions, mental health-related hospitalizations in the past year, serious systemic disorders, arterial hypotension, digestive tract diseases, recent major surgery, chronic/current use of certain medications and certain sleep medications, history of alcohol and drug abuse, current cancer diagnosis unless in remission for at least two years, allergies or sensitivity to GABA or placebo, or their ingredients, and MRI contraindications. **Table 1** summarizes our sample’s demographics.

**Table 1.** Sample characterization.

| **Variable** | **Statistic** |
| --- | --- |
| **Age, mean (SD)** | 68 (7.40) years |
| **Sex** |  |
| Male | 7 (21.2%) |
| Female | 26 (78.8%) |
| **Race** |  |
| Caucasian | 29 (87.9%) |
| African American | 3 (9.1%) |
| Hispanic | 1 (3.0%) |
| **Education Level** |  |
| High school | 7 (21.2%) |
| Two-year college | 8 (24.2%) |
| Four-year college | 8 (24.2%) |
| Master's degree | 7 (21.2%) |
| Doctorate's | 3 (9.1%) |
| **Marital Status** |  |
| Married | 19 (57.6%) |
| Other | 14 (42.4%) |

### Procedures

During the first visit (baseline visit), participants provided informed consent and underwent a general health assessment. Clinical pain, experimental pain and subjective sleep (e.g., PSQI) were also assessed. At the end of the visit, participants received an OURA© ring Generation 2, a smart device used primarily to monitor sleep. Participants were instructed to wear the ring continuously at home every night during sleep hours. They were also instructed to open the Oura mobile app each morning to enable the transfer, analysis, and upload of the ring’s data for subsequent access. After at least one month, participants returned for a follow-up visit to repeat the same assessments conducted during the baseline visit. Detailed information about these and other data can be found in the parent project’s publication^30^.

### Measures

#### Subjective Sleep Measure: The PSQI

This is a well-validated instrument used to assess sleep quality over a month using seven domains^8^: sleep quality, sleep latency, sleep duration, habitual sleep efficiency (ratio of total sleep time to bedtime), sleep disturbances, use of sleep-promoting medication, and daytime dysfunction. Each domain is rated from 0 to 3 (negative extreme). The sum is the PSQI total ranging from 0-21—a higher score indicates worse sleep quality.

#### Subjective Measures of the Impact of Sleep Quality on Function

**The Epworth Sleepiness Scale (ESS)**^24^. This is a self-administered questionnaire for assessing daytime sleepiness. Individuals rate their propensity to fall asleep in eight distinct situations, including while reading, watching TV, sitting inactively in a public place, etc. Each situation is scored from 0 (no chance of dozing) to 3 (high chance of dozing). Their sum is the total score, ranging from 0-24—a score of 10 or more is considered indicative of excessive daytime sleepiness.

**The Functional Outcomes of Sleep Questionnaire (FOSQ-10)**^11^. This self-administered instrument evaluates the impact of disorders of excessive sleepiness on activities of daily living. It has 10 questions assessing the ability to perform daily activities when feeling tired or sleepy. These activities include concentrating, remembering things, finishing a meal, working on a hobby, working around the house, operating a motor vehicle for short and long distances, driving or taking public transportation, and taking care of financial affairs and paperwork related to employed or volunteer work. Questions are scored from 1 (Yes, extreme difficulty) to 4 (No difficulty). The total score is the sum of the scores—higher scores indicate better functional status.

**Patient Reported Outcomes Measurement Information System (Short Forms) Sleep Related Impairment (SF PROMIS-SRI)**^44^. This instrument assesses self-reported perception of alertness, sleepiness and tiredness during usual waking hours, and the perceived functional impairments during wakefulness associated with sleep problems and impaired alertness, over 7 days. Raw scores are converted to T-scores (**PROMIS-SRI T-Score**) using the US population as a reference (mean/standard deviation=50/10)—higher T-scores indicate greater sleep impairment.

#### Objective Sleep Data: The OURA© Ring Gen2

Chosen for its compact design and ability to continuously track biometric data in real time over extended periods without frequent charging, it is suitable for older participants. The data collected were processed by the proprietary Oura Staging Algorithm to estimate sleep stages (light, deep, and REM) and generate several proprietary daily scores—ranging 0-100—namely the **Sleep Score**, **Sleep Efficiency Score**, **Restfulness Score**, **Sleep Latency Score**, **Sleep Timing Score**, as well as estimations of **Total Bedtime** (in hours), **Total Sleep Time** (in hours), and **Sleep Latency** (in minutes). These scores and estimates were used to derive Oura-based PSQI equivalents for our primary aim and averaged over all the days of usage for our second aim.

#### Oura-derived PSQI Equivalents

To compare subjective and objective sleep (our primary aim), we first calculated Oura-based equivalents for the PSQI responses and components where possible. Since the PSQI assesses sleep over the past 30 days, we used Oura scores and estimates recorded during the 30 days preceding the administration of the PSQI questionnaire to derive these equivalents. However, based on the available Oura data, only a subset of PSQI responses and components could be identified or calculated. The details of these equivalents are provided in **Table 2**.

**Table 2.** Details of the calculation of Oura’s equivalent items of the PSQI, used to test hypothesis 1.

| **PSQI item** | **Description of related Question, or Component** | **Values (Variable Type) Estimation** | **Oura equivalent** |
| --- | --- | --- | --- |
| PQSI1 | “During the past month, what time have you usually gone to bed at night?” | Hour (Hour) | Not available |
| PQSI3 | “During the past month, what time have you usually got up in the morning?” | Hour (Hour) | Not available |
| Total Bedtime | PQSI3 – PQSI1 | Hours (Continuous) | 1-month Average of Total Bedtime |
| PQSI2 | “During the past month, how long (in minutes) does it usually take you to fall asleep each night?” | Minutes (Continuous) | 1-month Average of Sleep Latency Score |
| PQSI4 | “During the past month, how many hours of actual sleep did you get at night? (This may be different than the number of hours you spent in bed)” | Hours (Continuous) | 1-month Average of Total Sleep Time |
| PQSI5a | “During the past month, how often have you had trouble sleeping because you: a) Cannot get to sleep within 30 minutes)” | Single choice (Ordinal, values shown below):<br>- Not during the past month (0)<br>- Less than once a week (1)<br>- Once or twice a week (2)<br>- Three or more times a week (3) | Occurrences of Sleep Latency Score > 30 min, then categorizing accordingly |
| C1 | Severity of Poor Overall Sleep Quality. This is PSQI9: “During the past month, how would you rate your sleep quality overall?” | Single choice (Ordinal, values shown below):<br>- Very good (0)<br>- Fairly good (1)<br>- Fairly bad (2)<br>- Very bad (3) | Sleep Score (0-100) categorized as:<br>[0, 55) → 3<br>[55, 70) → 2<br>[70, 85) → 1<br>[85, 100] → 0 |
| C2 | Severity of Longer Sleep Latency | 0, 1, 2, 3 (Ordinal)<br>Using PSQI2 and PSQI5a according to the PSQI’s scoring<sup>8</sup> | Using the Oura equivalents, according to the PSQI’s scoring<sup>8</sup> (see cells to the left) |
| C3 | Severity of Shorter Sleep Duration | 0, 1, 2, 3 (Ordinal)<br>Using PSQI4 according to the PSQI’s scoring<sup>8</sup> |  |
| C4 | Severity of Lower Efficiency | 0, 1, 2, 3 (Ordinal)<br>Using Total Bedtime and PSQI4<br>according to the PSQI's scoring<sup>8</sup> |  |
| C5 | Severity of Frequent Sleep Disturbances | 0, 1, 2, 3 (Ordinal)<br>Using PSQI5b-PSQI5j according<br>to the PSQI's scoring<sup>8</sup> | Since Oura equivalents of PSQI5b-PSQIj are not available, we used, as<br>a proxy, the Restfulness Score (0-100) categorized as:<br>[0, 70) → 3<br>[70, 85) → 2<br>[85, 100) → 1<br>100 → 0 |

Note. PQSI[X]: Answer to Question X of the PSQI. Higher component (C) values scores mean worse sleep quality. Averages were calculated excluding any missing entries.

To account for potential recall bias and the likelihood of forgetfulness regarding the most remote days when reporting sleep experiences in the PSQI, we recalculated the Oura equivalents as described in **Table 2**, but limiting the data to the most recent 25, 20, 15, or 10 days of ring usage.

#### Clinical Pain Measures

**Western Ontario and McMaster Universities Osteoarthritis Index - Pain (WOMAC-pain).** This subscale assessed pain experienced in the lower limbs during various activities^4^. Items were rated on a 5-point scale—higher scores, ranging from 0-20, indicate greater pain during activities.

**Short Form McGill Pain Questionnaire-Revised (SF-MPQ-2)**. This was used to measure the quality and the intensity of the current pain^28^. It comprised the SF-MPQ-2-Continuous pain, SF-MPQ-2-Intermittent pain, SF-MPQ-2-Neuropathic pain, and SF-MPQ-2-Affective experiences subscales. Each of 22 pain descriptors was rated on a 0 “no pain” to 10 “worst pain ever” scale within the past week, and a SF-MPQ-2-Total sum score was calculated for each subscale.

**Graded Chronic Pain Scale (GCPS)**. This 7-item questionnaire provides two subscales: pain intensity (GCPS-Intensity) and pain interference (GCPS-Interference) over the past 6 months^42^. For the GCPS-Intensity, participants were asked to rate on a 0 (“no pain”) to 10 (“pain as bad as could be”) numerical rating scale their current, average and worst pain. These were averaged and multiplied by 10 to yield a 0-100 score. Likewise, for the GCPS-Interference, participants were asked to rate on a 0 (“no inference”) to 10 (“unable to carry out activities”) scale how much pain has interfered with daily activities, recreational/social/family activities, and ability to work on average, over the past six months. These ratings were also averaged and multiplied by 10.

#### Experimental Pain Measures

Quantitative Sensory Testing (QST) was conducted on standardized anatomical sites and a region identified as painful by the participant using the TSA-II Neurosensory Analyzer and accompanying software (Medoc Ltd., Ramat Yishai, Israel), a TCS system (QST-Lab, https://www.qst-lab.eu/, France), and the AlgoMed computerized algometer (Medoc Ltd., https://www.medoc-web.com, Israel). More details can be found in our publication^30^.

**Vibration Detection Threshold**. Using the handheld VSA-3000 circular probe (contact tip=1.22 cm²) from the TSA-II Neurosensory Analyzer and accompanying software (Medoc Ltd., Ramat Yishai, Israel), a 100-Hz vibratory stimulus (ramping from 0μm at a 0.5μm/sec rate) was delivered to the thenar and painful site until the participant perceived the vibratory sensation. The average of the threshold across three trials, every 10 seconds, was calculated.

**Pressure Pain Threshold**. Using the AlgoMed computerized algometer (with a 10-mm rubber tip), pressure (kg/s rate) was applied to the right quadriceps muscle and a painful site—order randomized and counterbalanced—until the participant indicated that the sensation “first became painful.” The average threshold was calculated across three trials.

**Mechanical Temporal Summation**. A 300 g of pressure was applied to the thenar and the painful site using a nylon monofilament (TouchTest Sensory Evaluator 6.65)—order randomized and counterbalanced. Participants rated their pain after a single contact with the monofilament and after ten consecutive contacts at a 1 contact/s rate. The difference between these ratings was used to evaluate temporal summation of mechanical pain.

**Conditioned Pain Modulation (CPM)**. This was assessed to evaluate pain-inhibitory function. Heat pain (test stimulus) was applied to the left ventral forearm using the QST-Lab equipment with a T08 probe. Participants pressed a button when pain intensity reached a rating of 40/100. Participants then immersed their right hand in cold water (conditioning stimulus) for 60 seconds—temperature tailored to a pain rating of 40/100—and the procedure for the testing stimulus was repeated. CPM is defined as 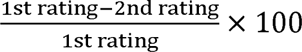.

### Statistical Analysis

Pearson’s partial correlations, controlling for age and sex, were used to assess the correlation between continuous variables and Spearman partial correlation for categorical variables. Pairwise deletion was applied, resulting in a variable sample size for each correlation.

To test our first hypothesis, we calculated the zero-order correlations between the PSQI items at follow-up and their corresponding Oura metrics from the preceding one-month period of ring usage, as outlined in **Table 2**. Since the PSQI captures self-reported sleep quality over the past month, it aligns temporally with the Oura data collected during the same period, making the follow-up PSQI more appropriate for this analysis than the baseline assessment. A one-tailed significance of 0.05 was applied, after Bonferroni correction for multiple comparisons.

For the second and third hypotheses, each variable of the first (the subjective sleep-related measures) and second column (the objective sleep-related measures) of **Table 3**, respectively, was correlated with each variable of the last two columns (the pain measures), controlling by age and sex (i.e., using partial correlations). A two-tailed statistical significance of 0.05 was applied, after Bonferroni correction for multiple comparisons.

**Table 3.** Variables used for the sleep-pain correlations (used for testing hypotheses 2 and 3).

| **Sleep Measures** |  | **Pain Measures** |  |
| --- | --- | --- | --- |
| *Subjective* | *Generated by OURA* | *Clinical* | *Experimental (QST)* |
| PSQI-Total Score | Sleep Score | WOMAC-Total | Vibratory Detection Threshold-Thenar |
| Epworth Total Score* | Sleep Efficiency Score | WOMAC-Pain | Vibratory Detection Threshold-Painful Site |
| FOSQ10 Total Score* | Restfulness Score | WOMAC-Stiffness | Mechanical Temporal Summation-Thenar |
| SF PROMIS-SRI T-Score* | Sleep Latency Score | WOMAC-Physical Function | Mechanical Temporal Summation-Painful Site |
|  | Sleep Timing Score | MPQ-Total | Pressure Pain-Threshold Quadriceps |
|  | Total Bedtime | MPQ-Continuous | Pressure Pain-Threshold Painful Site |
|  | Total Sleep Time | MPQ-Intermittent | Conditioned Pain Modulation |
|  | Sleep Latency | MPQ-Affective |  |
|  |  | MPQ-Neuropathic |  |
|  |  | GCPS-Intensity |  |
|  |  | GCPS-Interference |  |

Note. The measures generated by OURA, collected over several days between the baseline and follow-up visits, were averaged for this analysis. \*Subjective measures of the impact of sleep quality on function.

## Results

### Oura Ring Usage Summary

Within the one-month period preceding the PSQI administration at follow-up, from which Oura data was used for our primary aim, participants consistently wore the ring between 66.7% and 100% of the days. The mean usage was 92.8%, with a standard deviation of 11.7% and a median of 100%. More generally, across the entire study period, participants wore the Oura ring for 21 to 57 days, with a mean of 38.4 days, a standard deviation of 7.8 days, and a median of 39 days. These were the metrics used to compute the averages of Oura’s proprietary scores and estimates for our secondary aim. Further details on Oura ring usage relative to the baseline and follow-up visits are presented in **Figure 1**.

**Figure 1.**
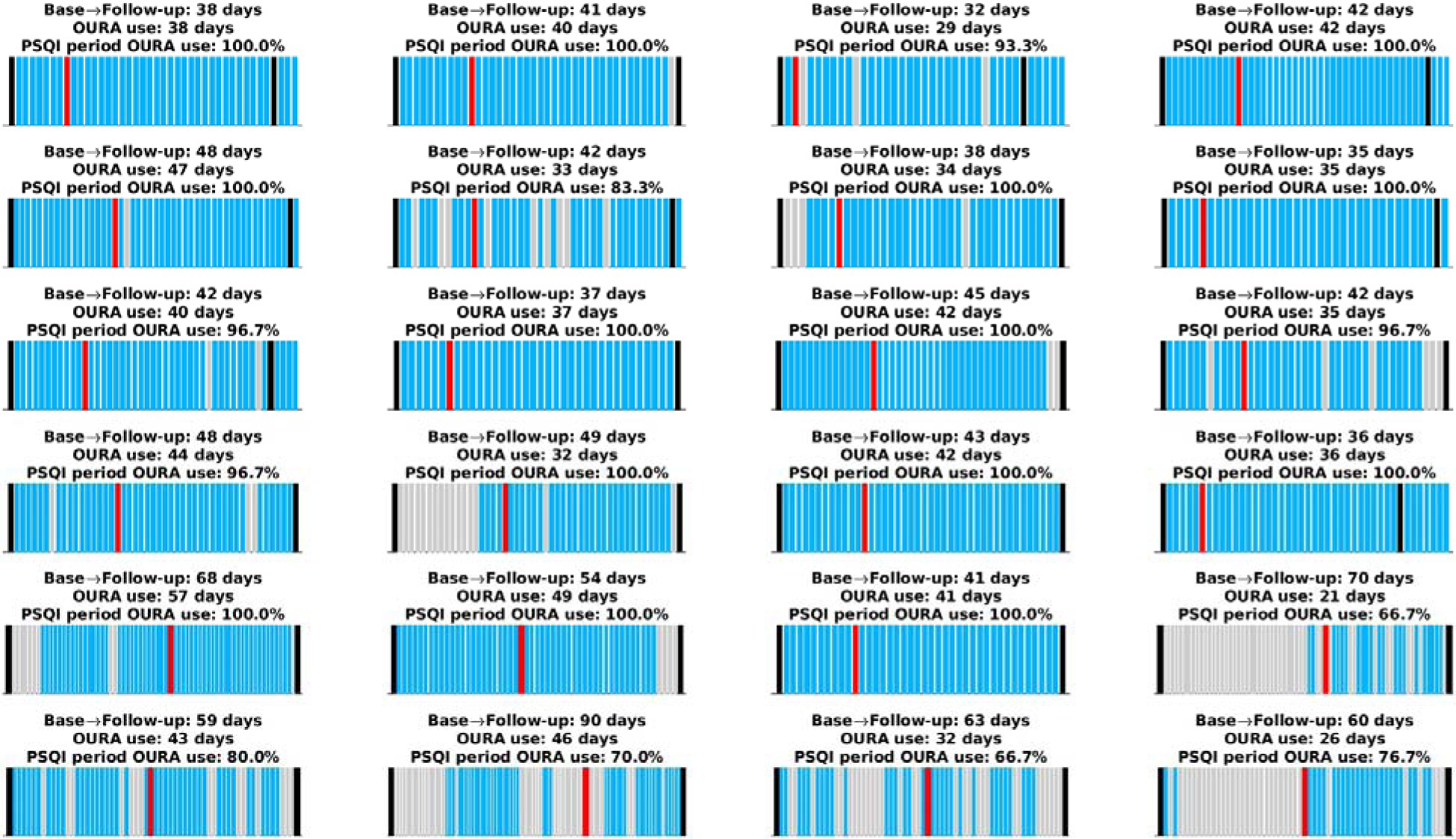
Oura Ring Usage for Each Participant. The x-axis spans the period from the time participants received the Oura ring until they returned it. Black lines indicate the baseline and follow-up visits. Blue bars represent days when the Oura ring was worn, while grey bars indicate days it was not worn. The red bar marks the start of the one-month period preceding the PSQI administration, which took place during the follow-up visit. Each subplot’s title provides i) the total number of days between the baseline and follow-up visits, ii) the total number of days the Oura ring was worn (used to compute averages for the second aim), and iii) the percentage of days the ring was worn within the one-month period preceding the PSQI administration (used to derive PSQI equivalents for the primary aim). Notably, some participants continued using the ring beyond the follow-up visit. Data from these additional days were only included in the averages used for the second aim.

### Comparison between Subjective and Objective Sleep

The correlation between the PSQI items and their 1-month Oura equivalents, introduced in **Table 2**, are shown in **Figure 2** (n=25 after pairwise deletion). A significant positive Pearson correlation was observed between the PSQI and the Oura equivalents for PSQI2 (r=0.49, p=0.051, though marginally, as it this seems to be driven by two outliers), PSQI4 (r=0.71, p<0.0005), and Total Bedtime (PSQI3−PSQI1; r=0.82, p<0.0005) and C3 (severity of longer sleep duration; r=0.62, p<0.003). There was a marginally significant positive correlation between the PSQI’s C1 (Severity of Poor Overall Sleep Quality; r=0.47, p<0.052). This is noteworthy because the correlation becomes significant when accounting for potential recall bias, specifically by limiting the Oura equivalents to any of the most recent 25, 20, 15, or 10 days of ring usage—see Figures S1-S4 in the Supplemental Materials. These figures also demonstrate that the remaining significant and non-significant results persist when accounting for potential recall bias.

**Figure 2.**
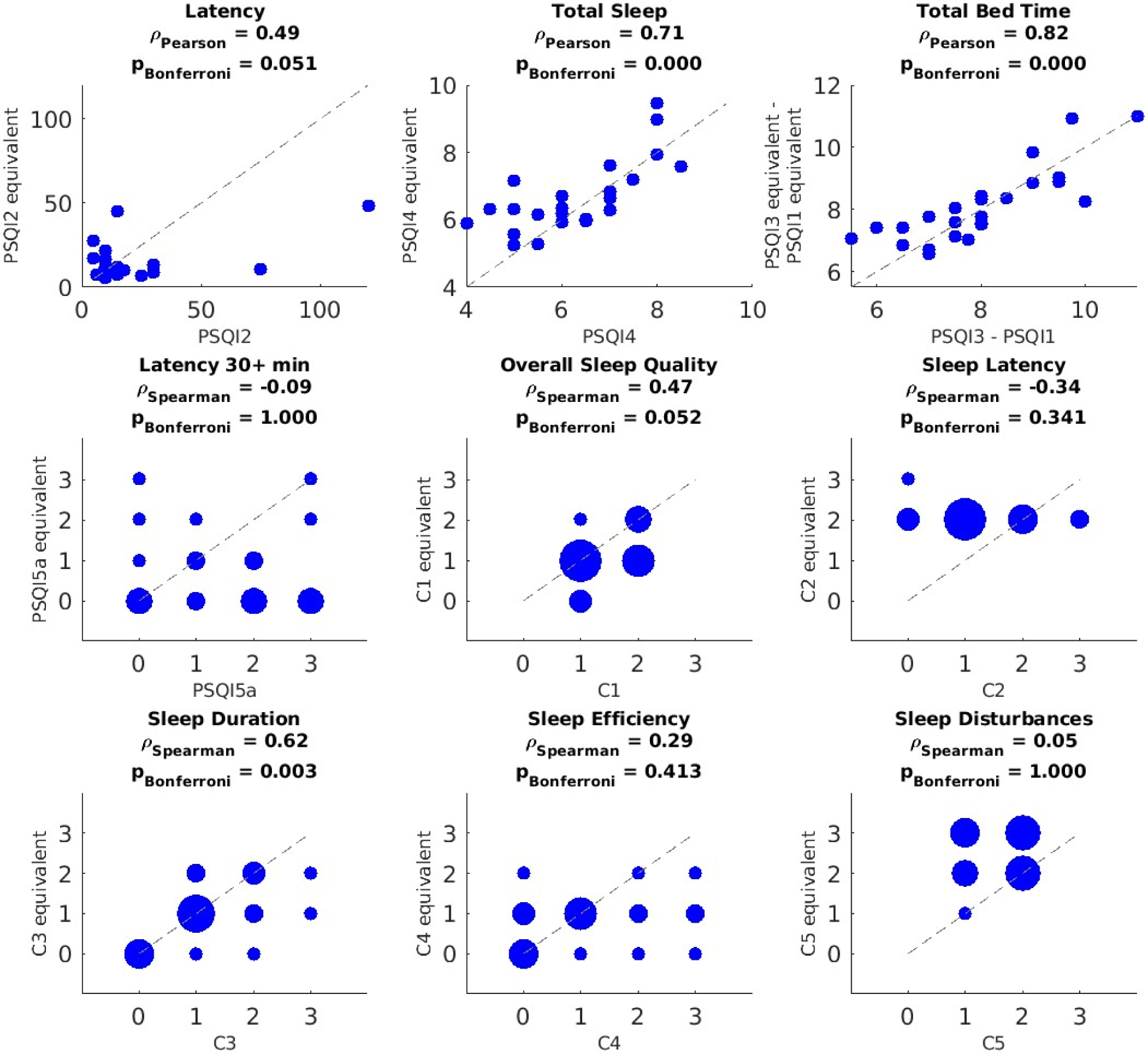
Correlation Between PSQI Measures and their Oura Equivalents Based on Data Recorded During the Month Preceding the PSQI Questionnaire. PSQI question or component (x axis) versus their Oura-derived equivalents (y axis). For the discrete variables (i.e., PSQI5a, C1-C5), blob size is proportional to the number participants in that value pair (bigger blobs in the y=x line means less discrepancy between the subjective measure and its objective equivalent). P-values were corrected using Bonferroni across questions and components independently. After pairwise deletion, the sample size used for these correlations was n=25.

### Comparison between Sleep and Pain

With a sample size determined by pairwise deletion, several significant correlations were observed after adjusting for multiple comparisons. PSQI-Total Score exhibited positive correlations with WOMAC-Pain (r = 0.53, p = 0.022, n=32), MPQ-Total (r = 0.50, p = 0.026, n=29), MPQ-Neuropathic (r = 0.45, p = 0.026, n=31), MPQ-Continuous (r = 0.48, p = 0.026, n=29), MPQ-Intermittent (r = 0.46, p = 0.026, n=31), and GCPS-Intensity (r = 0.45, p = 0.026, n=31). Additionally, PSQI’s C2 showed positive correlations with WOMAC-pain (r = 0.51, p = 0.009, n=32), MPQ-Total (r = 0.58, p = 0.008, n=29), MPQ-Neuropathic (r = 0.49, p = 0.011, n=31), MPQ-Intermittent (r = 0.55, p = 0.008, n=31), MPQ-Affective (r = 0.53, p = 0.008, n=32), GCPS-Intensity (r = 0.49, p = 0.012, n=31), and GCPS-Interference (r = 0.53, p = 0.008, n=31). Furthermore, the CPM showed a positive correlation with the average of Oura’s Sleep Latency (r = 0.626, p = 0.049, n=22). However, there were no significant associations between objective sleep measures and clinical pain measures.

## Discussion

Our study primarily aimed to evaluate the relationship between subjective and objective assessments of sleep quality in middle-aged and older adults with chronic musculoskeletal pain. Our findings suggest that the PSQI does not fully capture the complexity of sleep experiences in this population, particularly with respect to sleep latency and efficiency. Comparisons with sleep data acquired using the Oura ring suggested both alignment and discrepancies, underscoring the potential limitations of relying solely on self-reported measures in clinical and research settings.

Similar to studies using polysomnography (PSG) and actigraphy^5,12,20,34^, we observed correlations between self-reported sleep duration and Oura’s sleep duration. Our findings suggest that, despite inherent limitations such as recall bias^17^, subjective reports can offer valuable insights into sleep duration and bedtime in middle-aged and older adults with chronic pain. In our particular sample, the PSQI-Oura correlation observed for bedtime may be also enhanced by structured sleep habits often adopted by individuals managing chronic conditions—middle-aged and older adults with chronic pain frequently adopt consistent sleep schedules as part of self-management strategies aimed at reducing pain exacerbation and improving overall well-being^19,36^.

When considering the full month period, the PSQI-Oura correlation for overall sleep quality was marginal in our sample, becoming significant when increasing levels of recall bias were considered. A disagreement between objective and subjective measures of sleep quality over a longer period may be influenced by the interplay between physiological (e.g., hyperarousal), psychological (e.g., catastrophizing), and social factors, as proposed by the biopsychosocial model of pain^9,40^. Chronic pain is known to alter central nervous system processing and emotional states, which can distort the subjective perception of sleep quality^38^, exacerbating recall bias. For example, individuals with chronic pain may report poorer sleep quality due to heightened sensitivity to minor disturbances or difficulty dissociating pain from their sleep experience, even when objective indicators suggest otherwise.

Mirroring prior research^7^, our findings highlight discrepancies between subjective and objective sleep assessments and suggest that these discrepancies are not resolved by accounting for recall bias and may not depend on it. Specifically, the PSQI could not capture sleep latency (particularly on nights exceeding 30 minutes), efficiency and sleep disturbances. This discrepancy may stem from the inherent subjectivity of the PSQI, as individuals are often unaware of various factors that affect sleep scores, such as wake-after-sleep onset (WASO) or brief nighttime awakenings, which are, however, accurately detected by wearable devices.

For instance, while objective measures of WASO (Wake After Sleep Onset) were positively associated with the PSQI sleep disturbance component, the PSQI total score showed no correlation with objective sleep parameters^45^. Furthermore, chronic pain is associated with a heightened risk of sleep disturbances that may be underestimated through self-report alone^38^. Also, patients with chronic pain and insomnia often overestimate sleep latency compared to PSG^6^. Overall, the discrepancies between subjective and objective sleep assessments in our sample emphasize the need for cautious interpretation of self-reported sleep data and underscore the importance of integrating objective measures to achieve a more comprehensive evaluation of sleep quality in older adults with chronic pain.

Taken together, the findings related to our first hypothesis emphasize the benefits of a multimodal approach to sleep assessment, enabling a more comprehensive understanding of sleep quality by capturing both perceived and physiological aspects. These findings have important implications for clinical practice. Healthcare providers treating patients with chronic pain should consider incorporating objective sleep measurements, particularly in cases where poor sleep quality significantly impacts pain management and overall quality of life. Wearable devices like the Oura ring provide a non-invasive and reliable method for obtaining detailed sleep data, offering clinicians actionable insights to inform personalized treatment strategies.

Shifting focus to our second hypothesis, we found significant positive correlations between self-subjective overall sleep quality, as measured with the PSQI-Total Score, and various self-reported measures of pain severity. This finding aligns with the understanding that sleep and pain are closely interconnected and can influence each other^37–39^. Chronic pain can disrupt sleep, leading to poor sleep quality^15,22,25,33^, while poor sleep can, in turn, exacerbate pain perception and increase sensitivity to painful stimuli^4,27^. The positive correlation observed between the severity of longer sleep latency, assessed with the PSQI, and multiple pain measures suggests that difficulty falling asleep might be associated to the experience of pain in this population. This finding warrants further investigation to understand the specific mechanisms underlying this relationship, as well as its directionality.

Regarding our third hypothesis, no significant associations were found between pain and objective sleep data obtained from the Oura ring. However, given the significant relationship between pain and subjective sleep, this discrepancy invites further exploration of potential factors contributing to the divergence between subjective and objective sleep measures in individuals with chronic musculoskeletal pain. One possibility is that individuals with chronic pain may perceive sleep disturbances as more bothersome due to heightened pain sensitivity, potentially distorting their PSQI reports^10,43^. This distortion could weaken the alignment between subjective reports and the physiological sleep data captured by the Oura ring. While pain itself may be a contributing factor, this remains speculative and warrants further investigation. Future studies with adequate power should examine the moderating effect of pain on the relationship between objective and subjective sleep variables.

We observed a positive correlation between Conditioned Pain Modulation (CPM) and the 1-month average of Oura’s Sleep Latency. CPM reflects the body’s ability to regulate or reduce pain signals, relying on a complex interplay of neural pathways and neurotransmitters^35^. This correlation suggests that individuals with longer sleep latencies, indicating difficulty falling asleep, may experience alterations in their pain modulation capabilities. While preliminary, this finding underscores the need for further research to investigate the potential impact of sleep latency on pain modulation mechanisms.

Finally, psychological factors such as anxiety, depression, and stress, commonly comorbid with chronic pain^14,41^, can significantly impact both sleep quality and pain perception^1,26^. Conversely, positive encounters, such as meaningful social interactions or enjoyable daily experiences, may act as a buffer against these negative psychological influences^23^. These psychological factors are more likely to be reflected in self-reported measures like the PSQI than in objective sleep data.

### Limitations

The current study includes a small sample size, affecting power and demographic representativity, limiting the generalizability of the findings. Additionally, the study’s reliance on a single wearable device restricts the applicability of results to other devices with different algorithms or sensitivities. It is also essential to acknowledge the limitations of wearable technology in capturing the full spectrum of sleep experiences. Future research should explore larger, more diverse samples and consider other wearable technologies to assess the generalizability of these findings across different populations and devices. Longitudinal studies may also help determine how subjective and objective sleep measures vary over time and in response to different interventions.

### Conclusions

Our study highlights the need for integrating subjective and objective sleep assessments in individuals with chronic pain. While the PSQI provides insight into perceived sleep quality, it fails to capture key aspects like sleep latency and efficiency, contributing to discrepancies with objective measures. These differences may stem from recall bias, pain sensitivity, and biopsychosocial influences. The strong link between subjective sleep and pain severity, despite weak correlations with objective data, underscores the role of perception in sleep assessment. Incorporating wearable devices alongside self-reports provides a more comprehensive understanding of sleep disruptions in this cohort and supports the development of tailored interventions and pain management strategies to enhance overall well-being.

## Data Availability

Data will be available upon conclusion of the parent study and request to the authors

## Acknowledgments

We are grateful to our participants and study teams at the University of Florida (UF). We also extend our heartfelt thanks to Natalie J. Sawczuk and Barbara Gonzalez for their invaluable contributions.

## CRediT author statement

**Soamy Montesino-Goicolea**: Conceptualization, Methodology, Data curation, Formal analysis, Funding acquisition, Investigation, Methodology, Project administration, Visualization, Writing – original draft · **Pedro A. Valdes-Hernandez**: Formal analysis, Data curation, Methodology, Software, Visualization, Writing - Original Draft · **Olga Nin**: Resources, Writing – review & editing · **Eric C. Porges**: Supervision, Writing – review & editing · **Cameron Smith**: Resources, Writing – review & editing · **Yenisel Cruz-Almeida**: Funding acquisition, Project Administration, Resources, Supervision, Validation, Writing - review & editing, Supervision.

